# Predicting COVID-19 outbreak using open mobility data for minimal disruption on the country’s economy

**DOI:** 10.1101/2022.02.12.22270892

**Authors:** Héctor Morales-Fajardo, Jorge Rodríguez-Arce

**Affiliations:** Facultad de Ingeniería, Universidad Autónoma del Estado de México, Toluca, Mexico; Facultad de Medicina, Universidad Autónoma del Estado de México, Toluca, Mexico

**Author notes:** Email addresses:* (Héctor Morales-Fajardo); (Jorge Rodríguez-Arce).

**Keywords:** COVID-19, SARS-COV-2, prediction, mobility effects, mobility-based prediction

## Abstract

COVID-19 is an infectious disease caused by the SARS-COV-2 coronavirus, which was discovered in late 2019. Within a few months, COVID-19 was declared a global pandemic by the WHO. Several countries adopted social distancing measures, such as self-quarantine, workplace and mobility restrictions, reducing the probability of contact between non-infected and infected people. In general, these measures have a negative impact on low-income economies and small and medium businesses. During the outbreak, several predictive models have been proposed in order to assess the level of saturation that health services might have. Nevertheless, none of them considers information on the people’s mobility to assess the effectiveness of the social distancing policies. In this study, the authors propose a prediction method based on people’s open mobility data from Apple© and Google© databases to project potential scenarios and monitor case growth. The proposed method shows the importance of monitoring daily case increase for the first 4-6 weeks of the pandemic wave. Active monitoring is crucial to determine the reduction in mobility and proper actions. The results can contribute to health authorities for making timely decisions, preventing the spread of viruses while balancing the reduction of mobility with minimal disruption in people’s economies in future outbreaks.

## 1. Background

At the end of 2019, humanity has been challenged with a new pandemic caused by the novel corona virus identified as SARS-COV-2, leading to a global pandemic causing the disease called COVID or COVID-19. The outbreak was first identified in late December 2019, in the Wuhan province in China, when health authorities identified cases of atypical pneumonia of unknown origin and evidence of person-to-person transmission via droplets or direct contact [1] [2]. People were affected in different ways, developing mild to severe illness. Among the common symptoms were identified: fever, chills, muscle pain, headache, sore throat, cough, and shortness of breath (difficulty of breathing). After being infected with the virus, it can take up to 14 days for people to develop symptoms [3] [4]. According to World Health Organization (WHO), by the end of January 2020, China had more than 11,800 confirmed COVID-19 cases, as well as more than 250 deaths [5]. In a few months, the virus outbreak spread to countries in Asia, Europe, and America. On February 15th, 2020, WHO confirmed more than 40,000 cases in 28 countries. On March 11th, 2020, it also declared the outbreak as the Sixth Public Health Emergency and a global pandemic [6]. In order to avoid the increase on infections and overwhelm the health services of each country, WHO made the following recommendations:

- avoid close contact with sick people or with symptoms of COVID-19,
- avoid attending massive events as well as family gatherings,
- washing hands with soap and water for more than 20 seconds,
- cover mouth and nose with the elbow or a tissue when coughs or sneezes among others [1] [7].

Such contingencies measures that lead to greatest impact worldwide is social distancing such as travel and mobility restrictions as well as population confinement in their homes [8] [9].

Several countries adopted social distancing measures, such as self-quarantine, workplace and mobility restrictions in the public space, and facilities closures. Some of them in a voluntary manner by the citizens (such as Latin American countries) and others through strict mobility and travel restrictions (such as Spain, the United States, Italy or South Korea). All of these policies have the goal to reduce mobility in the public space, hence, reducing the probability of contact between non-infected and infected people, leading to avoidance of new community-transmitted cases [10][11][12][13]. For some countries, having the ability to measure population mobility variations is a challenge that must be solved in order to have data that helps predicting the spread of the disease. During this outbreak, predictive models have become a key tool for health authorities in each country to assess the level of saturation that health services might have. To date, no model has been able to accurately predict the evolution of future cases, however, depending on the variables involved, the number of people who could become infected and die, can be predicted.

Some authors have proposed different forecasting models. For example, a modified SEIR (Susceptible − Exposed − Infectious – Recovered) model to predict the COVID-19 outbreak was proposed by Löpez and Rodö [14]. They incorporated the effects of varying proportions of containment and fit data to quarantined populations accounting for the spread of infection during the outbreak. The proposed model predicts the number of infected individuals (undiagnosed) considering the different levels of isolation of the population, to evaluate the effects of the reduction of contacts. The model provides results similar to those obtained with more complex models. Li et. al [15] used the Gaussian distribution theory to propose a new model of COVID-19 transmission. The simulation results show that the curve of the proposed model is quite similar to the curve calculated with official data from different countries such as China, South Korea, Italy, and Iran. The model considers the virus incubation period, basic reproduction number, and daily infection number. A statistical forecast using time series models with open-data of confirmed cases, deaths, and recoveries was proposed by Petropoulos and Makridakis [16]. Using univariate time series models, and assuming that the data is accurate and that past patterns will continue to apply, forecast curves are obtained that could be used for decision-making and implementing control measures. The results obtained with these models show that forecast graphs of the COVID-19 outbreak can be calculated, however, none of them considers information on the people’s mobility to assess the effectiveness of the social restriction policies implemented in each country.

On the other hand, Oliver et. al [17] describe how mobile phone data could be used to design social distancing policies and delay the spread of the outbreak. The authors identified reasons why not all countries are able to use this information for predicting future cases. During the COVID-19 outbreak, some countries do not provide access to accurate information which might help to determine the mobility of their population and evaluate the effectiveness of social distancing policies. After WHO declared a global pandemic, Google© [18] and Apple© [19] released public mobility data from their platforms to help COVID-19 outbreak spread. Both companies decided to provide data about the people’s mobility patterns around the world based on information from mapping and GPS applications. Such information can be useful for health authorities as a basis for decision-making considering the volume of people who uses private vehicles, walks, or uses public transportation. Mexican government used this information to monitor people’s mobility in the public space [20].

As contribution, the authors propose a method for forecasting the outbreak using Google© and Apple©’s mobility trend reports. The authors envisions that this article can contribute to health authorities for making timely decisions, preventing the spread of viruses while balancing the reduction of mobility with minimal disruption in people’s economies in future outbreaks.

## 2. Technical Framework

In any infectious disease, there are a number of parameters to consider when countries are trying to control the outbreak. The most known parameter is vaccination, which allows to basically reduce to a manageable number of cases, ideally to zero. In cases with novel infectious agents such as the novel SARS-CoV-2 virus, which causes the disease called COVID-19, no vaccination yet exits, making public health officials being challenged to manage cases under control. For such cases, another parameter shall be used, called R which refers to the *effective reproduction number* [21] and, basically describes the way of measuring an infectious disease’s capacity to spread. In this study, the R number will be limited to understand outbreak spread [22], the use of it on the method and what it means for the average number of people that one infected person will pass the virus to [23].

The R number shall not be considered as a fixed parameter. Instead, it shall be seen as a parameter that can be affected by a range of factors [24], including not just how infectious a disease is but how it develops over time, how a population behaves in terms of mobility, containment and immunity. Immunity might be as a result of infection or vaccination.

A particular version of R is called R_0_ and is referred as the *basic reproduction number*, which is used when a new infection disease is discovered [22]. Another common use of R is called R_*e*_ or the *effective reproduction*. Ideally this number with vaccination is lower than 1.0 [25] [26].

In the case of SARS-CoV-2, R value varies from country to country, people’s behavior and population’s density: in urban areas, larger R_0_ values would be expected versus rural areas otherwise.

In order for any infectious disease such as the COVID-19 pandemic, is under control, R_0_ should be below 1. If R_0_ is equal to 1, then the outbreak is contained but never will disappear over the time. If R_0_ is greater than 1, then public health systems might suffer depending how lethal the infection agent is. Figure 1 shows different behaviors to an infection agent with just 10 identified cases in day 0. Over the time, depending upon the R_0_ value, cases might go down to zero (for R_0_ values less than 1) or might go up (for R_0_ values greater than 1). Hence the importance of controlling R_0_ values to manage outbreak spreads.

**Figure 1.**
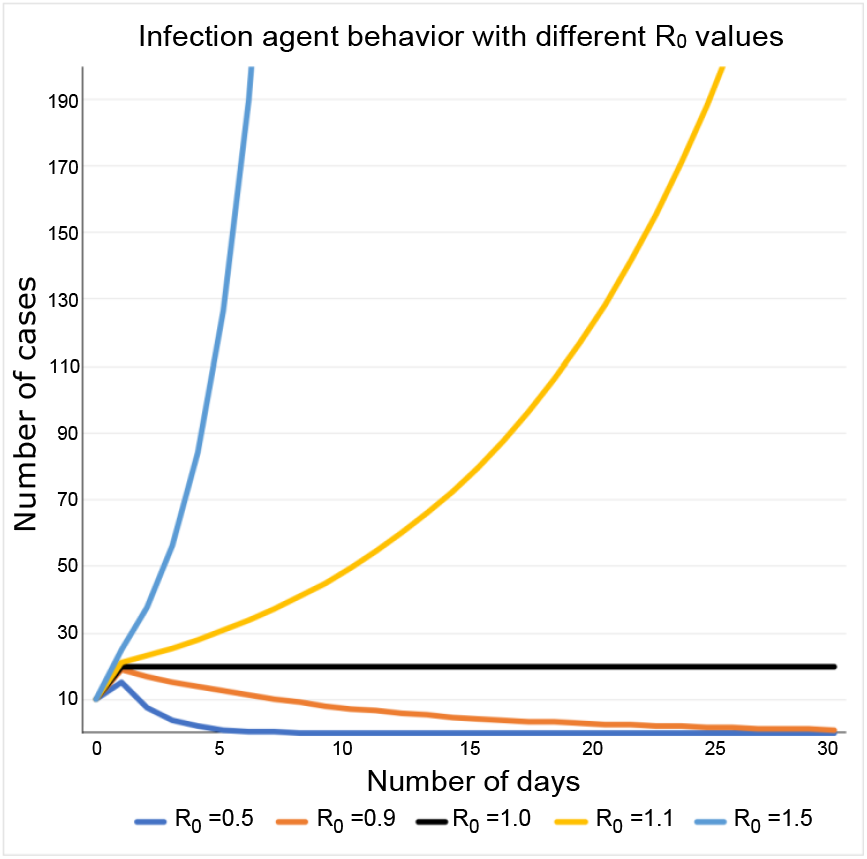
An infection agent such as SARS-COV-2 has a different behavior while reproduction number R_0_ varies.

According to WHO, one of the main strategies to contain the SARS-CoV-2 outbreak is via social distancing [27], meaning that people should maintain a safe separation from another individual, but also avoiding crowded places mainly in the public space. National state governments might adopt different measures to maintain social distances, some of them aiming public will and some others using enforcement agencies such as the police or national guards. The most important aspect is to keep people away from crowded places such as schools, public locations, office buildings, shopping centers and manufacturing floors.

Most of the national governments should keep a balance between public health, domestic macroeconomics and society well-being in general. Given the current outbreak conditions where there is no other treatment but keeping social distance, it results extremely important to make a decision on when the reduction in mobility should be in place and how deep this reduction should be applied in the public life. In this paper the authors propose a method to anticipate the growth of SARS-COV-2 in urban areas using open mobility data coming from Apple© iOS and Google© Android mobile phones. In addition, lessons learned from successful countries controlling the pandemic are discussed.

According to WHO there are four phases in a pandemic: Interpandemic phase, Alert phase, Pandemic phase and Transition phase, whereas the US Centers for Disease Control and Prevention, CDC, refers to 6 intervals: Investigation, Recognition, Initiation, Acceleration, Deceleration and Preparation. Figure 2 describes the pandemic curve of any infectious agent and both WHO and CDC phases or intervals. It is of our interest to understand the behavior of an outbreak in order to make the intervention measure more effective with the use of mobility data. Most of the current forecasting models [14][15][16] use the number of existing cases and make projections of future case growth using the expected outbreak scenario with and without interventions.

**Figure 2.**
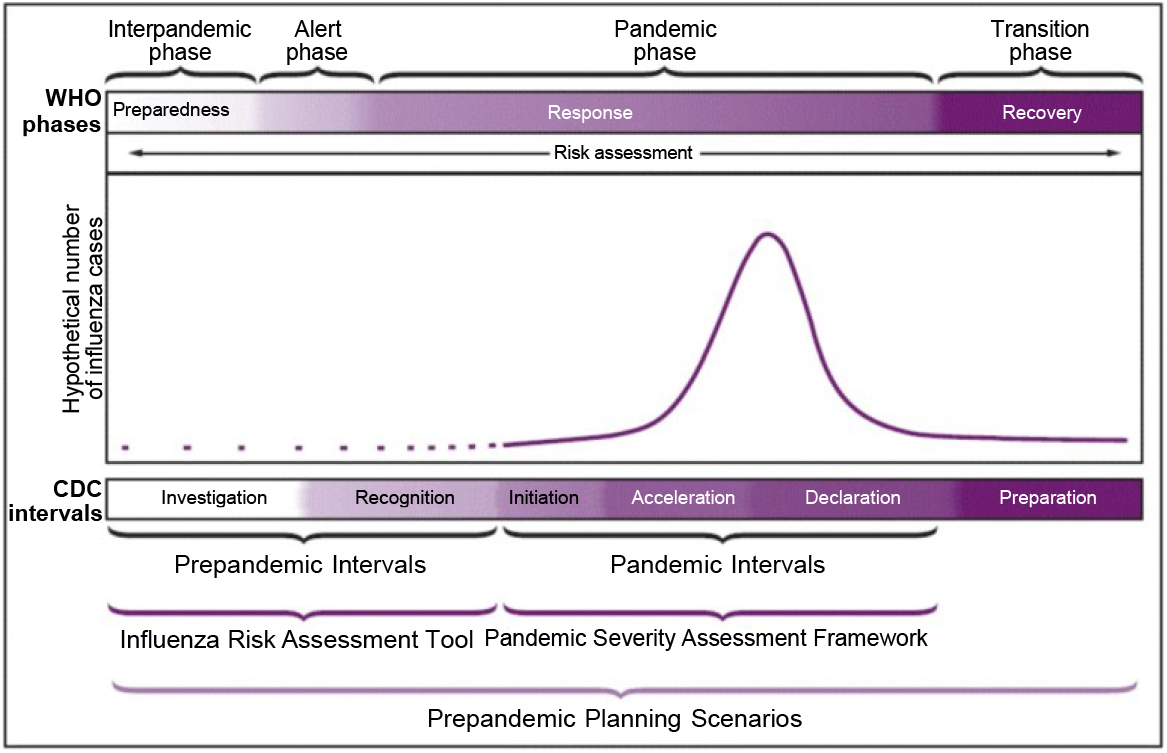
Preparedness and response framework for novel influenza A virus pandemics, with CDC intervals and WHO phases [28].

According to the Community Mitigation Guidelines to Prevent Pandemic Influenza [29] there shall be a risk assessment to understand the deepness of measures such as closure of schools or workplaces as well as the reduction of people’s mobility in the public space.

Table 1 shows the initial assessment in terms of transmissibility and clinical severity. As seen on the table, R_0_ greater than 1.8 means a moderate to high risk, as well as a case fatality rate greater than 10%. Hence the importance of predictive models that allow health officials to make decisions aiming to keepx R_0_ at low values.

**Table 1.** Initial assessment: scaled measures of influenza virus transmissibility and clinical severity (adapted from [30]).

| Measures of transmissibility and clinical severity | Scale |  |
| --- | --- | --- |
|  | Low to moderate | Moderate to high |
| **Transmissibility** |  |  |
| Secondary attack rate, household | ≤20% | >20% |
| Attack rate, school or university | ≤30% | >30% |
| Attack rate, workplace or community | ≤20% | >20% |
| R0, basic reproductive number | 1.0-1.7 | ≥1.8 |
| Underlying population immunity | Some underlying population immunity | Little to no underlying population immunity |
| Emergency department or other outpatient visits for influenza-like illness | <10% | ≥10% |
| Virologic characterization | Generic markers for transmissibility absent | Genetic markers for transmissibility present |
| Animal models, transmission studies | Less efficient or similar to seasonal influenza | More efficient than seasonal influenza |
| **Clinical severity** |  |  |
| Upper bound of case-fatality ratio | <1% | ≥1% |
| Upper bound of case-hospitalization ratio | <10% | ≥10% |
| Deaths-hospitalizations ratio | <10% | ≥10% |
| Virologic characterization | Genetic markers for virulence absent | Genetic markers for virulence present |
| Animal models, evaluation of morbidity and mortality | Less virulent or similar to seasonal influenza | More virulent than seasonal influenza |

For a specific country or location, the future case growth can be expressed as a function of its population mobility parameter called ***m***_0_, and its basic reproduction number ***R***_0_.

Table 2 shows that different values of ***R***_0_ and the impact in future case growth with only 10 cases in day 1 and assuming ***R***_0_ is not affected. According to pandemic mitigation this action would be referred as a scenario with no intervention [31]. Equation 1 shows the case growth based on ***R***_0_:

**Table 2.** Case growth of any given infectious agent with A) R_0_ = 1.1 B) R_0_ = 1.5 and C) R_0_ = 1.8.

| Day | Daily Cases | R0 | Day | Daily Cases | R0 | Day | Daily Cases | R0 |
| --- | --- | --- | --- | --- | --- | --- | --- | --- |
| 1 | 10 | 1.10 | 1 | 10 | 1.50 | 1 | 10 | 1.80 |
| 2 | 11 | 1.10 | 2 | 15 | 1.50 | 2 | 18 | 1.80 |
| 3 | 12 | 1.10 | 3 | 23 | 1.50 | 3 | 32 | 1.80 |
| 4 | 13 | 1.10 | 4 | 34 | 1.50 | 4 | 58 | 1.80 |
| 5 | 15 | 1.10 | 5 | 51 | 1.50 | 5 | 105 | 1.80 |
| 6 | 16 | 1.10 | 6 | 76 | 1.50 | 6 | 189 | 1.80 |
| 7 | 18 | 1.10 | 7 | 114 | 1.50 | 7 | 340 | 1.80 |
| 8 | 19 | 1.10 | 8 | 171 | 1.50 | 8 | 612 | 1.80 |
| 9 | 21 | 1.10 | 9 | 256 | 1.50 | 9 | 1102 | 1.80 |
| 10 | 24 | 1.10 | 10 | 384 | 1.50 | 10 | 1984 | 1.80 |
| 11 | 26 | 1.10 | 11 | 577 | 1.50 | 11 | 3570 | 1.80 |
| 12 | 29 | 1.10 | 12 | 865 | 1.50 | 12 | 6427 | 1.80 |
| 13 | 31 | 1.10 | 13 | 1297 | 1.50 | 13 | 11568 | 1.80 |
| 14 | 35 | 1.10 | 14 | 1946 | 1.50 | 14 | 20823 | 1.80 |
| 15 | 38 | 1.10 | 15 | 2919 | 1.50 | 15 | 37481 | 1.80 |

A) B) C)

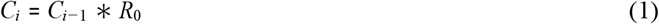

where:

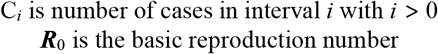

The importance of mobility parameter ***m***_0_ in pandemic mitigation for COVID-19 is key to make the cases below the health system capacity ***h***_*c*_ shown in Figure 3. Herein, the main contribution of this method is on finding the proper mobility ***m***_0_ parameter via public mobility data such that cases never overpasses ***h***_*c*_. Ideally ***h***_*c*_ value shall be below maximum capacity to avoid saturation of public health system or within a toleration threshold given that health capacity will not be overpassed.

**Figure 3.**
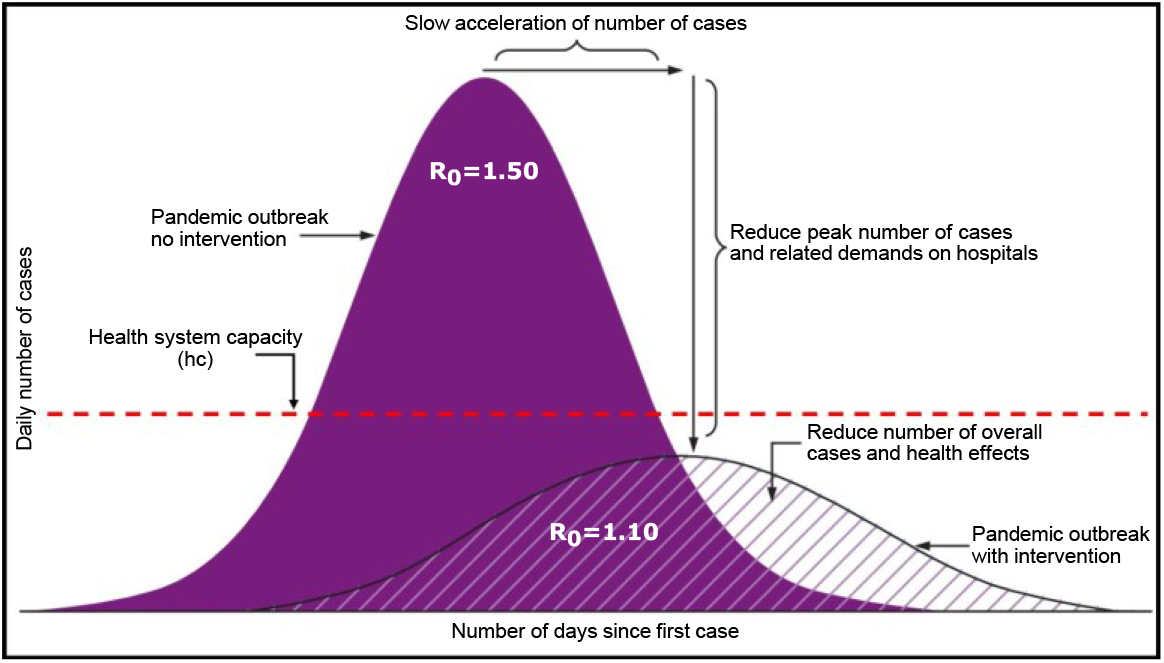
Adapted from: CDC. Interim pre-pandemic planning guidance: community strategy for pandemic influenza mitigation in the UnitedStates— early, targeted, layered use of nonpharmaceutical interventions and capping to Health System Capacity [31].

## 3. Model and Data sets

As explained in section 2, the main goal of this research is to find how mobility impacts in future case growth and, eventually, be able to use mobility parameter ***m***_0_ as the controlling factor to reduce SARS-COV-2 transmission, in a form of case future growth reduction, given that there is no current vaccination available.

In general terms, the *case growth* can be represented as a function of the *number of cases in the period i* as ***C***_*i*_ and its *basic reproduction number* ***R***_0_ as described in Equation 1. If *i* = 1 then ***C***_0_ is the initial case of the pandemic. In our case, this is the first documented case and this will indicate the first day of the epidemic curve.

When ***R***_0_ is greater than 1 and there is no intervention [31], then the number of cases tends to grow exponentially until the outbreak reaches its natural peak.

Herein the importance of ***R***_0_ to be controlled with some mechanism of intervention. In this case, the use of ***m***_0_ is proposed and for that matter, the use of mobility data provided by mobile applications such as Google© and Apple© data.

In this study, ***R***_*t*_ means the *temporal reproduction number*. Consequently ***R***_0_ can then be represented as a function of ***m***_0_ parameter to smoothen the exponential growth of daily cases. Equation 2 shows this expression:

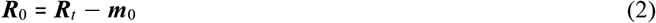

where:

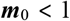

to make

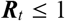

Equation 1 can be rewritten as:

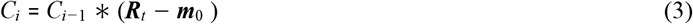

***R***_0_ and ***m***_0_ parameters are calculated in subsequent sections.

### 3.1. Data sets

Data sets for COVID-19 cases were taken from the Journal “Our world in data” [32]. Mobility data was taken from Google© [18] and Apple© [19] public mobility data sets and associated mobility data to May 15th, 2020 for Apple© and to May 9th, 2020 for Google©. Eight countries’ data were analyzed based on global representation from different regions of the globe and based on existing available public mobility data:

- Australia
- Brazil
- Italy
- Mexico
- South Korea
- Spain
- United Arab Emirates (UAE)
- United States (US)

The analysis period was limited to the dates that Google© and Apple© provided mobility data and up to the date mentioned above (first date reported by Apple© was January 13th, 2020 and Google© was February 15th, 2020). At least it was able to analyze the first 70 days of the pandemic wave in each country.

In order to make a fair comparison between the countries’ growth and mobility, dates were not considered, instead number of days since the first official case was documented. For the purpose of the research it will be referred to the initiation phase or as long as acceleration phase, according to WHO [31], has been identified, instead of a particular date.

### 3.2. Determination of m_0_

In order to determine a useful index it was necessary to analyze current mobility data provided by Google© and Apple©.

For Apple© [19], mobility data is provided using the maps application and the directions users are looking for. Apple© also disclaims that mobility provided it is a representation of the total population, but not the overall by any mean. The mobility data is reported to be 100 for the first date (January 13th, 2020), meaning that a greater value than 100 means more mobility whereas a value lesser than 100 means otherwise. Apple© reports three types of mobility data, driving, transit and walking.

For Google© [18], mobility data is reported as a positive percent of change from baseline when there is more mobility, whereas negative means less mobility. First reported date is February 15h, 2020. Google© reports six types of mobility data: retail and recreation, grocery and pharmacy, parks, transit stations, workplaces and residential.

According to Google’s methodology [33], mobility trends are displayed by country or region and how visitors in categorized locations change compared to the baseline date mentioned above. Each high-level category contains many types of places, so it was needed to understand and select those categories that would impact on mobility for potential infection outbreaks. For the purpose of our research and based on Google© methodology to use mobility data, only the percent of change associated to retail, recreation, grocery, pharmacy, parks, transit and workplaces will be used.

Based on both Apple © and Google © mobility data sets, a new data set was created for each country for a number of days (greater than 70 days) where mobility information was available. Each of the mobility data from both sources were averaged to find a single daily mobility score per country. This new data set shows the combined index that will be used to evaluate the case growth. This mobility score will be referred as the Combined Mobility Index (CMI), which will be used to determine the ***m***_0_ parameter.

The CMI data indicates how many people in a particular location is reducing their mobility in the public space. Table 3 shows the CMI of different countries, an indication of low mobility reductions is when people reduces their mobility less than 25% (represented by red dot), mid mobility reductions represented by a yellow dot is in the range of 26% to 50%, and larger mobility reductions represented by a green dot is when reduction is larger than 51%.

**Table 3.** Snippet of the Combined Mobility Index per each country. Table shows first 14 days of pandemic wave per each country.

| Combined Mobility Index (CMI) |  |  |  |  |  |  |  |  |
| --- | --- | --- | --- | --- | --- | --- | --- | --- |
| Day since first case | US | Mexico | Brazil | Italy | Spain | UAE | Australia | South Korea |
| 1 | ● -0.61 | ● 3.59 | ● -4.34 | ● 14.66 | ● 42.04 | ● 1.23 | ● -5.93 | ● 8.28 |
| 2 | ● 0.37 | ● 15.76 | ● 1.30 | ● 27.38 | ● 13.09 | ● 1.06 | ● -15.20 | ● 4.87 |
| 3 | ● 4.01 | ● 17.92 | ● 6.98 | ● 11.94 | ● 9.41 | ● -2.00 | ● -2.04 | ● 6.40 |
| 4 | ● 21.48 | ● -2.23 | ● 5.48 | ● 2.50 | ● 10.11 | ● 7.47 | ● 1.59 | ● 18.90 |
| 5 | ● 20.80 | ● 0.06 | ● -10.39 | ● 3.46 | ● 11.30 | ● 11.47 | ● 1.07 | ● 18.03 |
| 6 | ● -13.81 | ● 0.61 | ● 2.02 | ● 3.59 | ● 16.68 | ● 6.52 | ● 1.89 | ● 23.12 |
| 7 | ● -4.18 | ● 1.06 | ● 3.04 | ● 7.02 | ● 38.18 | ● -1.55 | ● 3.40 | ● 12.55 |
| 8 | ● -2.47 | ● 3.64 | ● 3.27 | ● 21.65 | ● 42.17 | ● 0.28 | ● -10.08 | ● 1.12 |
| 9 | ● -1.96 | ● 13.88 | ● 3.25 | ● 35.62 | ● 10.98 | ● -3.46 | ● -5.06 | ● 0.56 |
| 10 | ● 2.76 | ● 16.78 | ● 10.38 | ● 16.31 | ● 9.51 | ● 1.31 | ● -3.12 | ● -2.57 |
| 11 | ● 20.70 | ● -0.95 | ● 7.18 | ● 7.33 | ● 10.05 | ● 12.13 | ● 0.04 | ● 7.19 |
| 12 | ● 22.18 | ● -15.63 | ● -11.16 | ● 11.80 | ● 14.11 | ● 16.71 | ● 0.11 | ● 8.12 |
| 13 | ● -15.38 | ● 0.30 | ● -1.07 | ● 15.08 | ● 23.61 | ● 5.35 | ● 2.08 | ● -14.55 |
| 14 | ● -4.08 | ● 2.03 | ● 4.18 | ● 22.13 | ● 30.91 | ● 1.22 | ● 7.03 | ● -20.76 |

Using CMI allows to provide an understanding on people’s mobility trends and how this might impact in future case growth. From Table 3, Italy and Spain show a low decrease in people’s mobility, indicated by the red dots, whereas Australia and South Korea show some yellow dots, indicating an early trend in diminishing people’s mobility. Contrasting to Mexico and Brazil, which in fact showed some decrease in people’s mobility, is less than the one shown by Australia and South Korea.

### 3.3. Determination of R_t_

An important element to understand any pandemic wave behavior is the reproduction number, either basic ***R***_0_ or effective ***R***_*e*_ as explained in Section 2. Given the complexity of determining such a number, the daily growth based on daily confirmed cases was used to obtain the ***R***_0_ that will be used to find the expected ***R***_*t*_. Important to mention that confirmed cases might not be dated when symptoms appeared but instead, the date they were reported. Still, it is valid for the analysis and for the model. ***R***_*t*_ is calculated using Equation 4:

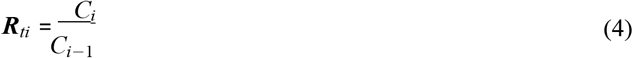

Equation 4 shows the calculation of daily reproduction number, hence the *i* index based on case number *i*.

It is also known that the SARS-COV-2 has an incubation period no longer than 14 days [34] and according to WHO’s recommendations [27], this period of quarantine is enough to keep infected individuals safer to not spread the virus.

A rolling average, also known as moving average or running average, is the unweighted mean of the last *n* values of a data set [35]. The rolling average is used to understand the trends of a data set, in this case to understand the trend of the ***R***_*t*_ and the ***CMI***. Based on these facts, ***R***_*t*_ and ***CMI*** in 14-days rolling average per each country was calculated. Table 4 shows the first 14-day rolling averages of these pandemic wave calculations per country.

**Table 4.** Snippet of Combined Mobility Index and Temporal Reproduction Number in 14-day rolling average per each country for the first 28 days of the pandemic wave.

| 14-day Rolling Average |  |  |  |  |  |  |  |  |  |  |  |  |  |  |  |  |
| --- | --- | --- | --- | --- | --- | --- | --- | --- | --- | --- | --- | --- | --- | --- | --- | --- |
| Day since first case | US |  | Mexico |  | Brazil |  | Italy |  | Spain |  | UAE |  | Australia |  | South Korea |  |
|  | CMI | Rt | CMI | Rt | CMI | Rt | CMI | Rt | CMI | Rt | CMI | Rt | CMI | Rt | CMI | Rt |
| 14 | 3.56 | 1.26 | 4.06 | 1.25 | 1.44 | 1.36 | 14.32 | 1.00 | 20.15 | 1.08 | 4.12 | 1.28 | -1.73 | 1.34 | 5.09 | 1.27 |
| 15 | 3.45 | 1.24 | 4.04 | 1.26 | 2.11 | 1.36 | 15.33 | 1.00 | 19.36 | 1.07 | 4.03 | 1.26 | -2.28 | 1.32 | 3.04 | 1.25 |
| 16 | 3.29 | 1.24 | 3.87 | 1.24 | 2.20 | 1.40 | 15.77 | 1.00 | 19.51 | 1.07 | 3.99 | 1.27 | -1.37 | 1.10 | 0.96 | 1.26 |
| 17 | 3.16 | 1.25 | 3.54 | 1.21 | 1.97 | 1.43 | 16.23 | 1.00 | 19.75 | 1.07 | 4.26 | 1.27 | -1.08 | 1.10 | -1.42 | 1.27 |
| 18 | 2.99 | 1.25 | 3.19 | 1.21 | 1.15 | 1.45 | 16.90 | 1.00 | 20.19 | 1.07 | 4.66 | 1.06 | -0.95 | 1.10 | -4.59 | 1.29 |
| 19 | 3.24 | 1.18 | 1.24 | 1.25 | 0.31 | 1.40 | 17.53 | 1.00 | 20.67 | 1.07 | 5.05 | 1.06 | -0.61 | 1.10 | -7.01 | 1.22 |
| 20 | 3.48 | 1.18 | 0.05 | 1.26 | -0.93 | 1.45 | 18.35 | 1.00 | 21.02 | 1.07 | 5.08 | 1.06 | 0.37 | 1.07 | -10.09 | 1.22 |
| 21 | 3.61 | 1.07 | -1.72 | 1.28 | -2.80 | 1.46 | 19.12 | 1.00 | 20.91 | 1.07 | 5.49 | 1.04 | 1.47 | 1.06 | -12.54 | 1.19 |
| 22 | 3.87 | 1.08 | -3.98 | 1.30 | -5.04 | 1.48 | 19.33 | 1.00 | 20.69 | 1.07 | 5.69 | 1.05 | 2.29 | 1.04 | -13.98 | 1.17 |
| 23 | 4.27 | 1.08 | -7.12 | 1.32 | -7.62 | 1.47 | 18.77 | 1.33 | 21.37 | 1.07 | 6.22 | 1.05 | 2.91 | 1.02 | -15.55 | 1.17 |
| 24 | 4.87 | 1.08 | -10.91 | 1.34 | -10.89 | 1.39 | 18.19 | 1.59 | 21.78 | 1.00 | 6.41 | 1.05 | 3.47 | 1.02 | -16.57 | 1.17 |
| 25 | 4.96 | 1.07 | -14.16 | 1.33 | -14.43 | 1.37 | 17.52 | 1.64 | 21.95 | 1.04 | 6.31 | 1.05 | 3.85 | 1.02 | -18.18 | 1.17 |
| 26 | 4.78 | 1.06 | -16.27 | 1.34 | -17.09 | 1.39 | 15.82 | 1.69 | 22.08 | 1.18 | 5.74 | 1.05 | 4.38 | 1.02 | -18.97 | 1.12 |
| 27 | 6.01 | 1.05 | -19.74 | 1.35 | -20.37 | 1.35 | 13.70 | 1.72 | 21.94 | 1.24 | 5.58 | 1.04 | 5.02 | 1.01 | -19.15 | 1.07 |
| 28 | 6.60 | 1.02 | -23.47 | 1.32 | -24.38 | 1.37 | 11.23 | 1.74 | 22.09 | 1.32 | 5.51 | 1.05 | 5.65 | 1.01 | -18.93 | 1.05 |
| 29 | 7.03 | 1.02 | -27.32 | 1.30 | -28.57 | 1.35 | 8.49 | 1.79 | 22.12 | 1.36 | 5.49 | 1.05 | 7.04 | 1.03 | -18.34 | 1.05 |
| 30 | 7.43 | 1.02 | -31.79 | 1.27 | -32.51 | 1.32 | 5.10 | 1.81 | 21.89 | 1.39 | 5.34 | 1.04 | 7.54 | 1.03 | -17.23 | 1.05 |
| 31 | 7.67 | 1.02 | -36.14 | 1.25 | -36.49 | 1.30 | 2.16 | 1.83 | 21.59 | 1.44 | 5.10 | 1.04 | 7.85 | 1.03 | -16.05 | 1.08 |
| 32 | 7.72 | 1.02 | -39.56 | 1.24 | -39.93 | 1.30 | -0.13 | 1.87 | 21.19 | 1.47 | 4.29 | 1.04 | 8.12 | 1.03 | -15.35 | 1.11 |
| 33 | 7.59 | 1.11 | -41.33 | 1.21 | -42.61 | 1.29 | -2.16 | 1.88 | 20.74 | 1.50 | 3.39 | 1.07 | 8.24 | 1.03 | -15.86 | 1.17 |
| 34 | 8.38 | 1.11 | -43.93 | 1.20 | -45.45 | 1.25 | -4.83 | 1.90 | 20.28 | 1.52 | 2.85 | 1.07 | 7.91 | 1.04 | -16.37 | 1.26 |
| 35 | 8.76 | 1.11 | -46.08 | 1.19 | -47.73 | 1.24 | -7.80 | 1.91 | 19.73 | 1.56 | 2.38 | 1.08 | 7.77 | 1.03 | -16.52 | 1.31 |
| 36 | 8.98 | 1.14 | -48.10 | 1.17 | -49.61 | 1.24 | -11.08 | 1.93 | 19.33 | 1.59 | 1.79 | 1.07 | 8.39 | 1.04 | -17.65 | 1.32 |
| 37 | 9.13 | 1.14 | -49.95 | 1.16 | -51.26 | 1.22 | -14.72 | 1.61 | 18.82 | 1.62 | 1.04 | 1.07 | 8.53 | 1.04 | -18.12 | 1.33 |
| 38 | 8.91 | 1.14 | -51.25 | 1.16 | -52.58 | 1.20 | -17.96 | 1.37 | 18.15 | 1.65 | 0.36 | 1.09 | 8.52 | 1.05 | -19.05 | 1.35 |
| 39 | 8.83 | 1.14 | -52.24 | 1.15 | -53.55 | 1.18 | -21.21 | 1.34 | 17.38 | 1.65 | -0.55 | 1.09 | 8.39 | 1.06 | -20.83 | 1.38 |
| 40 | 9.04 | 1.14 | -53.18 | 1.15 | -54.39 | 1.17 | -24.44 | 1.31 | 15.86 | 1.54 | -1.36 | 1.10 | 8.08 | 1.08 | -23.30 | 1.40 |
| 41 | 8.97 | 1.15 | -53.93 | 1.15 | -54.86 | 1.15 | -28.24 | 1.29 | 12.57 | 1.50 | -1.75 | 1.08 | 7.75 | 1.10 | -25.05 | 1.43 |

When contrasting rolling averages of CMI shown in table 4 with the previous daily values of CMI shown in table 3, it results evident to find how much progress countries might be having to decrease people’s mobility in the public space. A clear example of early measures is shown in South Korea, which consistently decreases to reach 25% in 40 days. In contrast, Mexico and Brazil shown decreases in the order of 50% in the same period of time. However, the use of ***R***_*t*_ is particularly useful to identify how quickly the pandemic wave might be evolving. US, Italy and Spain are very interesting examples of low reduction on mobility with a quick increase on ***R***_*t*_.

## 4. Results

Based on the numbers for ***CMI*** and ***R***_*t*_ shown in Table 4, the behavior of these parameters was analyzed over the 6 periods of 14-days (with the exception of Mexico and Brazil, which spans for 78 and 81 days respectively). A 14-day rolling average of both parameters was calculated in order to understand the trend, if any, and finally a Pearson’s correlation between these two data sets of parameters on a per country-basis was done, in order to identify how much mobility, would impact the reproduction number.

In a Pearson’s correlation *r* statistic [36], when one variable increases the other variable has a tendency to also increase. This is called a positive correlation. A negative correlation indicates when one variable increases the other variable has a tendency to decrease. Finally, there is no correlation when one variable increases, the other variable does not tend to either increase or decrease [37].

Cohen [38] and Evans [36] proposed empirical classifications of interpreting correlation strength by using Pearson *r* statistic. Cohen recommended that 0.10 to 0.30 be interpreted as a weak correlation, 0.30 to 0.50 as a moderate correlation and greater than 0.50 as a strong correlation. According to Evans’ classification, less than 0.20 is very weak, 0.20 to 0.39 is weak, 0.40 to 0.59 is moderate, 0.60 to 0.79 is strong and 0.80 or greater is a very strong correlation for the absolute value of r [39].

A statistically significant correlation does not necessarily mean that the strength of the correlation is strong. In order to determine whether the correlation between variables is significant, it is necessary to compare the p-value to a significance level. The *p-value* shows the probability that this strength may occur by chance [40]. A significance level, denoted as α or alpha, of 0.05 is generally acceptable. This means that the risk of concluding that a correlation exists when no correlation exists is 5%.

Based on this analysis, using Evans’ classification and *p-value*<0.05, countries were classified in three main sets:

- Countries with Pearson values greater of 0.60 or with a strong to very strong correlation.
- Countries with Pearson values between 0.40 to 0.59 or with moderate correlation.
- Countries with Pearson values less than 0.39 or with weak or very weak correlation.

All of them under the assumption that *p-value* has a high statistical significance with *p-value*<0.05. As described, correlation strength is not enough, but is necessary to determine the statistical significance. The results of these new calculations are also shown in Table 5.

**Table 5.**
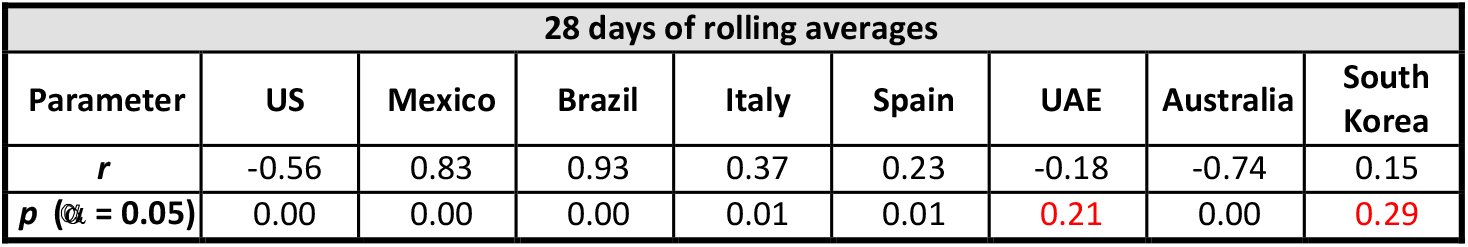
Pearson correlation and p-value for 28-day rolling averages of ***CMI*** and ***R***_*t*_.

As shown in table 5, countries might have a positive or negative correlation, low or high correlation strength, but the most important value to make an initial decision if the proposed model would work is the statistical significance *p-value*. If p >0.05, then the correlation has not statistical significance, meaning the correlation between data sets is not significant, so can’t lead to any decision.

From table 5 can be seen that Brazil and Mexico have a strong correlation, meaning that their mobility reduction and reproduction rates were strong correlated, whereas Italy and Spain have weak correlations. Impact of cases growth for Italy and Spain is expected and could be seen with the correspondent impact on health systems overloads. The particular case of United States where there is a negative correlation is a clear indication that case might be out of control due to mobility increase. For the cases of South Korea and UAE, the authors do not recommend using this method because there is no statistical significance between the CMI and Rt values.

## 5. Discussion

The main goal of the proposed method is to find ***m***_0_ that contributes to control number of cases, which is a function of ***R***_*t*_ subject to the health capacity system ***h***_*c*_.

Maximum number of cases will determine the potential number of people requiring hospitalization services, hence controlling ***R***_*t*_ via ***m***_0_ is fundamental.

The proposed method is described below:

1. Given initial number of cases, determine ***R***_*ti*_ as described in Section 3. Authors suggest to have at least two full 14-day periods of confirmed cases in order to provide an approximation of ***R***_*t*_. Remember that for the purpose of the calculations ***R***_*t*_ = ***R***_*e*_.
2. Obtain daily ***CIM*** as described in Section 3. For the purpose of this research, Apple© and Google© Mobility Data was used, but any mobility data can be used.
3. A valid Pearson correlation of ***CIM*** and ***R***_*t*_ pairs should be obtained. Authors suggest to calculate a p-value, such that p-value < 0.05 provides a valid statistical significance for Pearson correlation. The stronger Pearson correlation and the p-value <0.05, the better the validation of **CIM and Rt** will be.
4. Obtain 7-day rolling averages of ***CIM*** and ***R***_*t*_. For example, if the data set has 30 days of cases, then, 23 7-day rolling averages can be obtained.
5. Using the last ***CIM*** and ***Rt*** rolling averages as the seed data, projections can be done, where ***m***_0_ value could be changed to make ***R***_*e*_<1.

From Section 4, future case growth depends on initial number of cases but also in two determining factors:

- Number of confirmed cases in the second 14-day period, ideally in less than 40 days.
- Daily Effective Reproduction Number for the same period.

For the purposes of the discussion, ***R***_*t*_ (temporal reproduction number) is in fact the ***R***_*e*_ number for the pandemic wave, meaning that the *effective reproduction number* will be considered as the actual daily case growth. The bigger ***R***_*e*_ were in that second period, a greater mobility reduction would be needed to decelerate future case growth.

However, the biggest challenge found in most of the predictive models relies on the fact that confirmed cases are cumulative and some countries may change their method to identify cases depending upon the pandemic behavior. Counting cases and expecting changes on cases is not scalable nor practical to analyze.

Also, none of the models suggests ways to control the behavior of the pandemic, so this is the opportunity this model is intended to address.

In order to describe the model from a practical perspective, actual cases from one country were chosen and its data is shown in Table 6. Number of daily confirmed cases from *“Our World in Data”* journal database [32] were used to calculate daily ***R***_*e*_ using Equation 4. Daily ***CIM*** was calculated as well for the same period of time using Google© [18] and Apple© [19] mobility data. Selected country had a Pearson correlation of 0.58, moderate according to Evan’s model [36] with p-value of 0.001 with α = 0.05, in a period of 30 days.

**Table 6.**
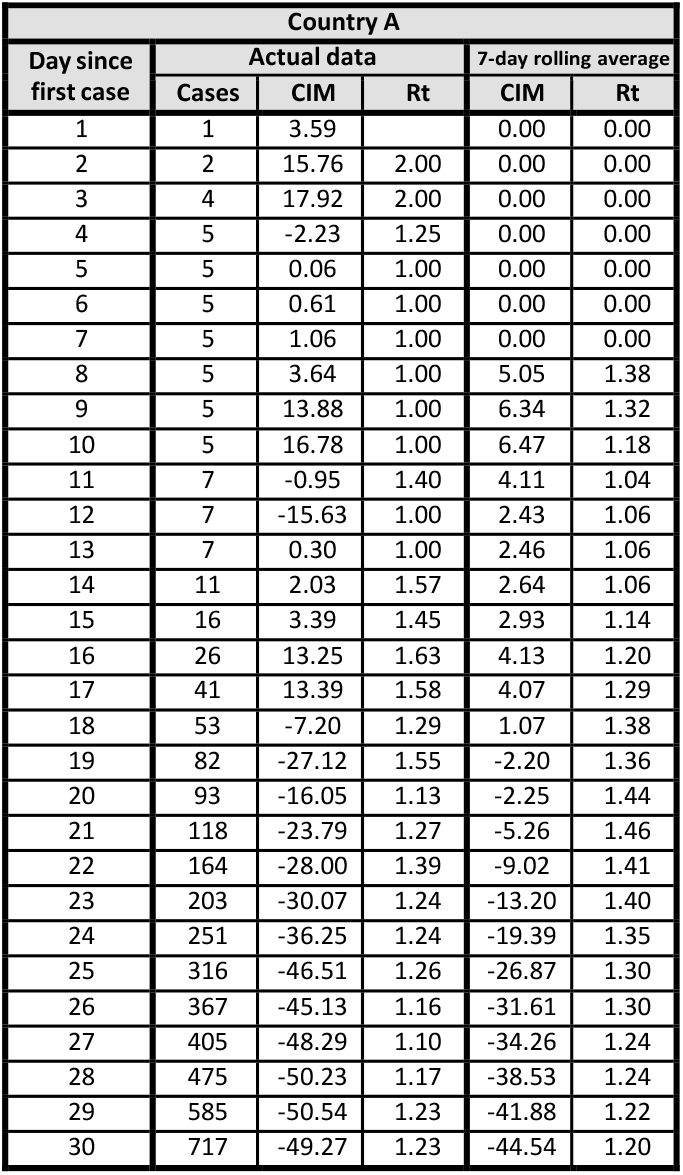
First 30 days of actual reported cases, Combined Mobility Index and Temporal Reproduction Number.

| Country A |  |  |  |  |  |
| --- | --- | --- | --- | --- | --- |
| Day since first case | Actual data |  |  | 7-day rolling average |  |
|  | Cases | CIM | R<sub>t</sub> | CIM | R<sub>t</sub> |
| 1 | 1 | 3.59 |  | 0.00 | 0.00 |
| 2 | 2 | 15.76 | 2.00 | 0.00 | 0.00 |
| 3 | 4 | 17.92 | 2.00 | 0.00 | 0.00 |
| 4 | 5 | -2.23 | 1.25 | 0.00 | 0.00 |
| 5 | 5 | 0.06 | 1.00 | 0.00 | 0.00 |
| 6 | 5 | 0.61 | 1.00 | 0.00 | 0.00 |
| 7 | 5 | 1.06 | 1.00 | 0.00 | 0.00 |
| 8 | 5 | 3.64 | 1.00 | 5.05 | 1.38 |
| 9 | 5 | 13.88 | 1.00 | 6.34 | 1.32 |
| 10 | 5 | 16.78 | 1.00 | 6.47 | 1.18 |
| 11 | 7 | -0.95 | 1.40 | 4.11 | 1.04 |
| 12 | 7 | -15.63 | 1.00 | 2.43 | 1.06 |
| 13 | 7 | 0.30 | 1.00 | 2.46 | 1.06 |
| 14 | 11 | 2.03 | 1.57 | 2.64 | 1.06 |
| 15 | 16 | 3.39 | 1.45 | 2.93 | 1.14 |
| 16 | 26 | 13.25 | 1.63 | 4.13 | 1.20 |
| 17 | 41 | 13.39 | 1.58 | 4.07 | 1.29 |
| 18 | 53 | -7.20 | 1.29 | 1.07 | 1.38 |
| 19 | 82 | -27.12 | 1.55 | -2.20 | 1.36 |
| 20 | 93 | -16.05 | 1.13 | -2.25 | 1.44 |
| 21 | 118 | -23.79 | 1.27 | -5.26 | 1.46 |
| 22 | 164 | -28.00 | 1.39 | -9.02 | 1.41 |
| 23 | 203 | -30.07 | 1.24 | -13.20 | 1.40 |
| 24 | 251 | -36.25 | 1.24 | -19.39 | 1.35 |
| 25 | 316 | -46.51 | 1.26 | -26.87 | 1.30 |
| 26 | 367 | -45.13 | 1.16 | -31.61 | 1.30 |
| 27 | 405 | -48.29 | 1.10 | -34.26 | 1.24 |
| 28 | 475 | -50.23 | 1.17 | -38.53 | 1.24 |
| 29 | 585 | -50.54 | 1.23 | -41.88 | 1.22 |
| 30 | 717 | -49.27 | 1.23 | -44.54 | 1.20 |

Using this data, 7-day rolling averages were calculated for ***R***_*t*_ and ***CIM*** data of both parameters was used as seed for projections, also shown in Table 6. Variable parameter for projections is a value of ***m***_0_ given that resulting ***R***_*t*_ is less or equal to 1, hence number of cases will tend to stall or decrease over time. For this country, three scenarios were projected using the proposed method:

- **Scenario 1:** Light constant mobility reductions (***CIM***=***-1***) until reach ***R***_*e*_<1. Peak number of cases is about 22,400 cases with a total reduction of mobility of -81.5% to reach ***R***_*t*_=0.99 and keep going down. The disadvantage of this approach is a hard reduction in mobility of more than 80% for more than 60 days. This scenario is inviable for low-income economies and small and medium business companies. Figure 4 shows total number of cases and behavior over the time.

**Figure 4.**
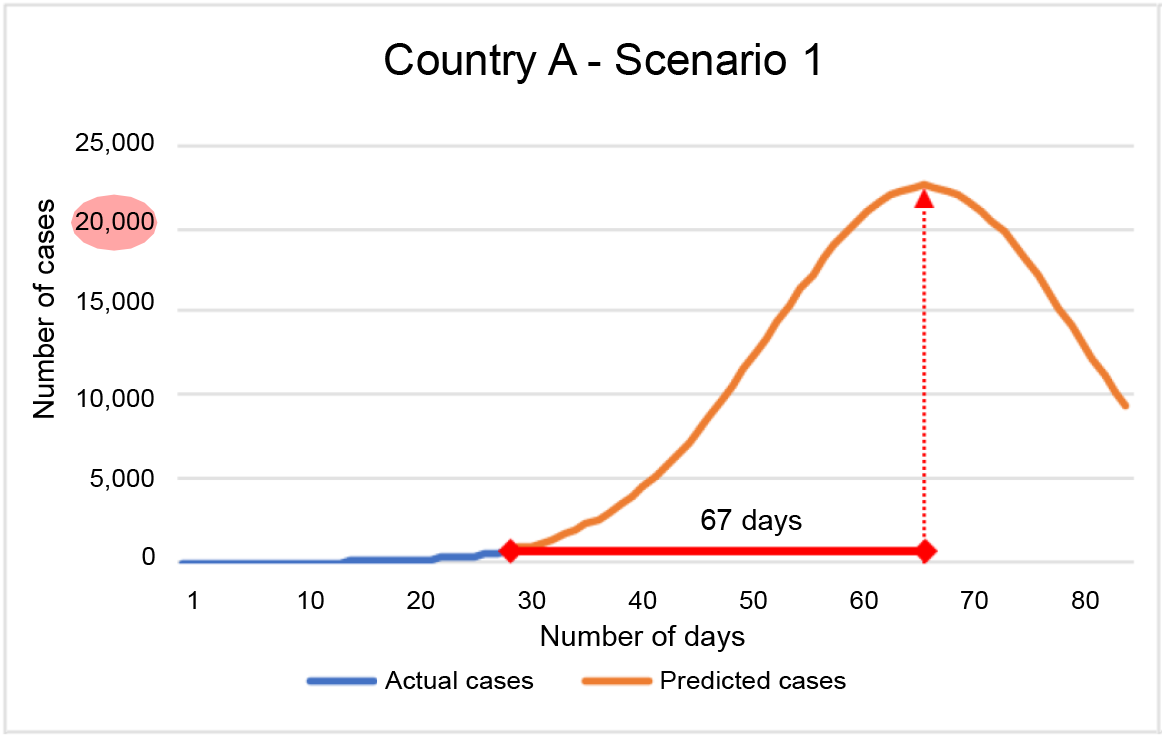
Country A, Scenario 1 - Peak number of cases were around 22,000 cases on day 66 since first documented case or 36 days after the last know data. Number of cases would decrease due to constant mobility measures, which makes this an inviable scenario.
- **Scenario 2:** Moderate reductions (***CIM***=***-3***) on mobility for the first 2 weeks and then relaxed measures (***CIM***>***0***) moving forward. In low income economies, a balance between a lockdown and public health is needed, hence, authors suggest a 2-week projections as viable. Peak number of cases is about 2,100 cases and ***R***_*t*_=0.99 before mobility measures were relaxed below -80%, where number of cases started to increase over 5,000 cases. Figure 5 shows total number of cases and behavior over the time. This scenario illustrates a lack of monitoring in mobility causing the virus to spread again, with increased number of cases.

**Figure 5.**
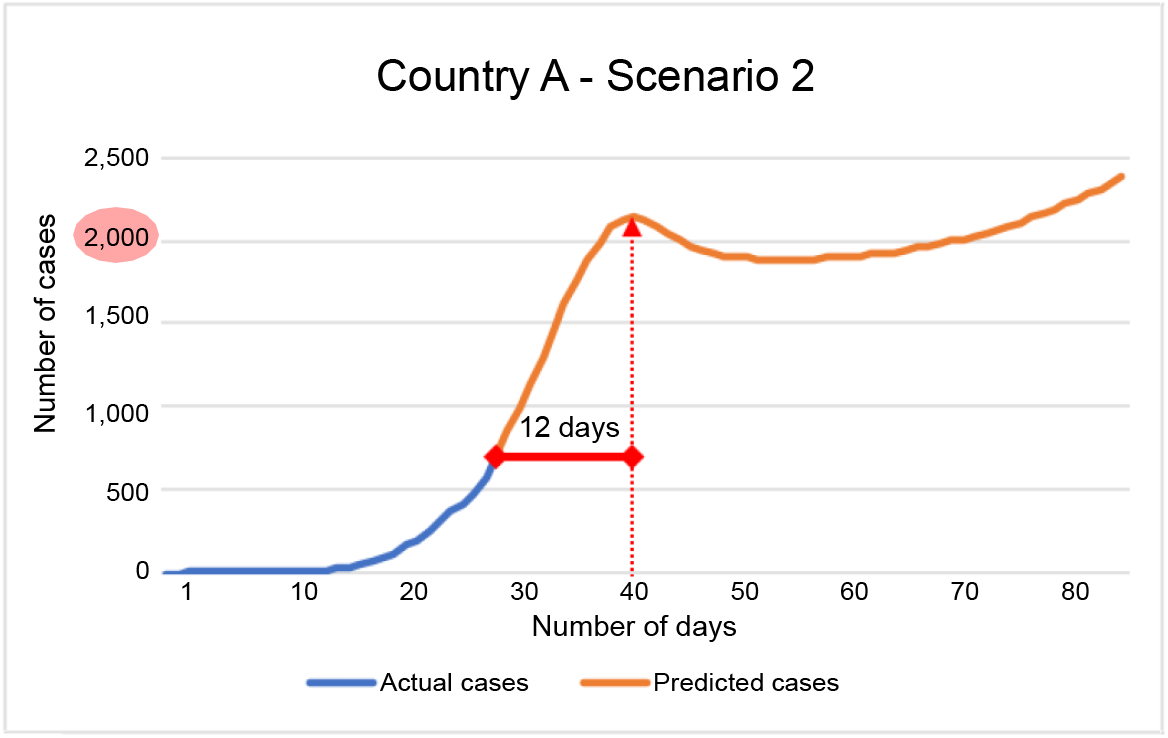
Country A, Scenario 2 - Peak number of cases were around 2,100 cases on day 42 since first documented case or 12 days after the last know data. Number of cases would increase due to mobility measures relaxation after day 56 and on.
- **Scenario 3:** Strong reductions on mobility for the first 5 days (***CIM***=***-5***), moderate measures for the next 5 days (***CIM***=***-3***), and relaxed measures after 2 weeks. After the tenth day, ***R***_*t*_=0.98, peak number of cases is about 1,400 with a total reduction of mobility of -79.56%. On the projected eleventh day, people can start moving, hence a constant surveillance of ***R***_*t*_ is needed to avoid cases growth as a result of increased mobility. Figure 6 shows total number of cases and behavior over the time.

**Figure 6.**
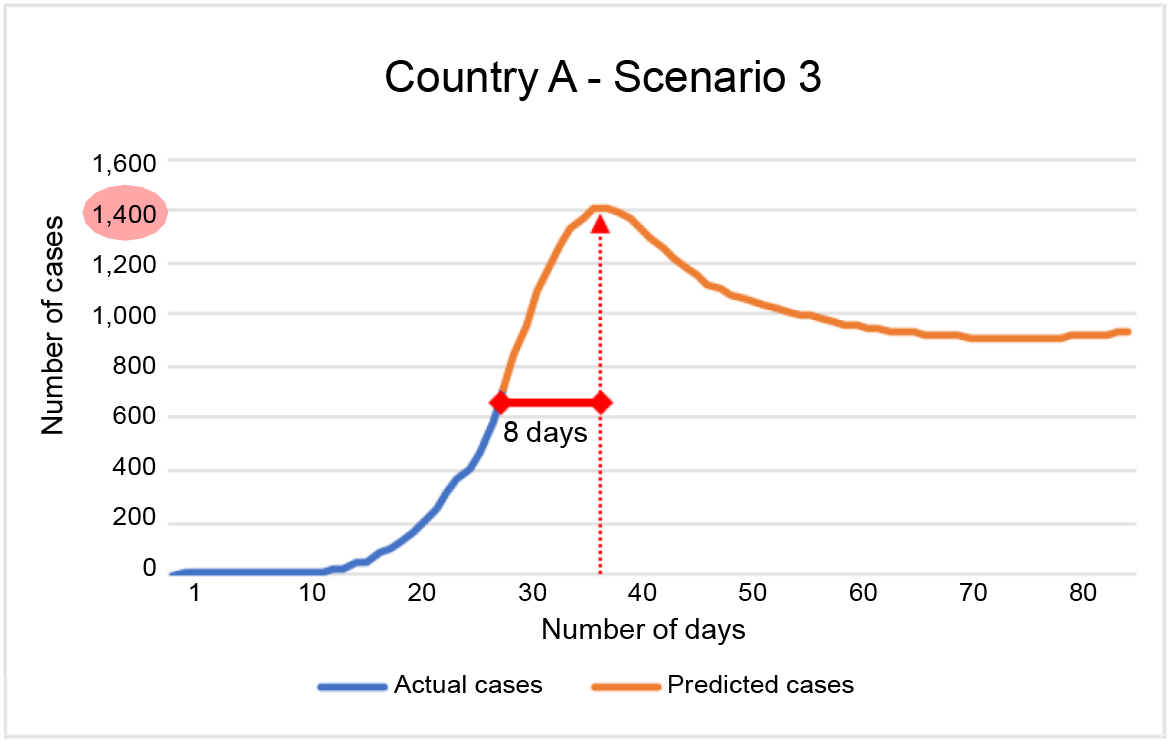
Country A, Scenario 3 - Peak number of cases were around 1,400 cases on day 38 since first documented case or 8 days after the last know data. Number of cases would increase due to mobility measures relaxation.

Looking into real data, a similar scenario 1 corresponds to a similar behavior show by Italy and Spain, both had apparently a good control in number of cases however, both had a large ***R***_*e*_ for that period, more specifically a large ***R***_*e*_ 14-day rolling average in the first 4 weeks. In order to achieve pandemic control, it was necessary to go to levels of 80% of mobility reduction over a period of 100 days in order to reach ***R***_*e*_<***1***, making economy practically to stop. This measure results inviable from economic and social perspectives plus the risk of crashing the health system as occurred unfortunately in both countries. Figure 7 shows the actual number of cases and needed ***CIM*** to decelerate pandemic wave.

**Figure 7.**
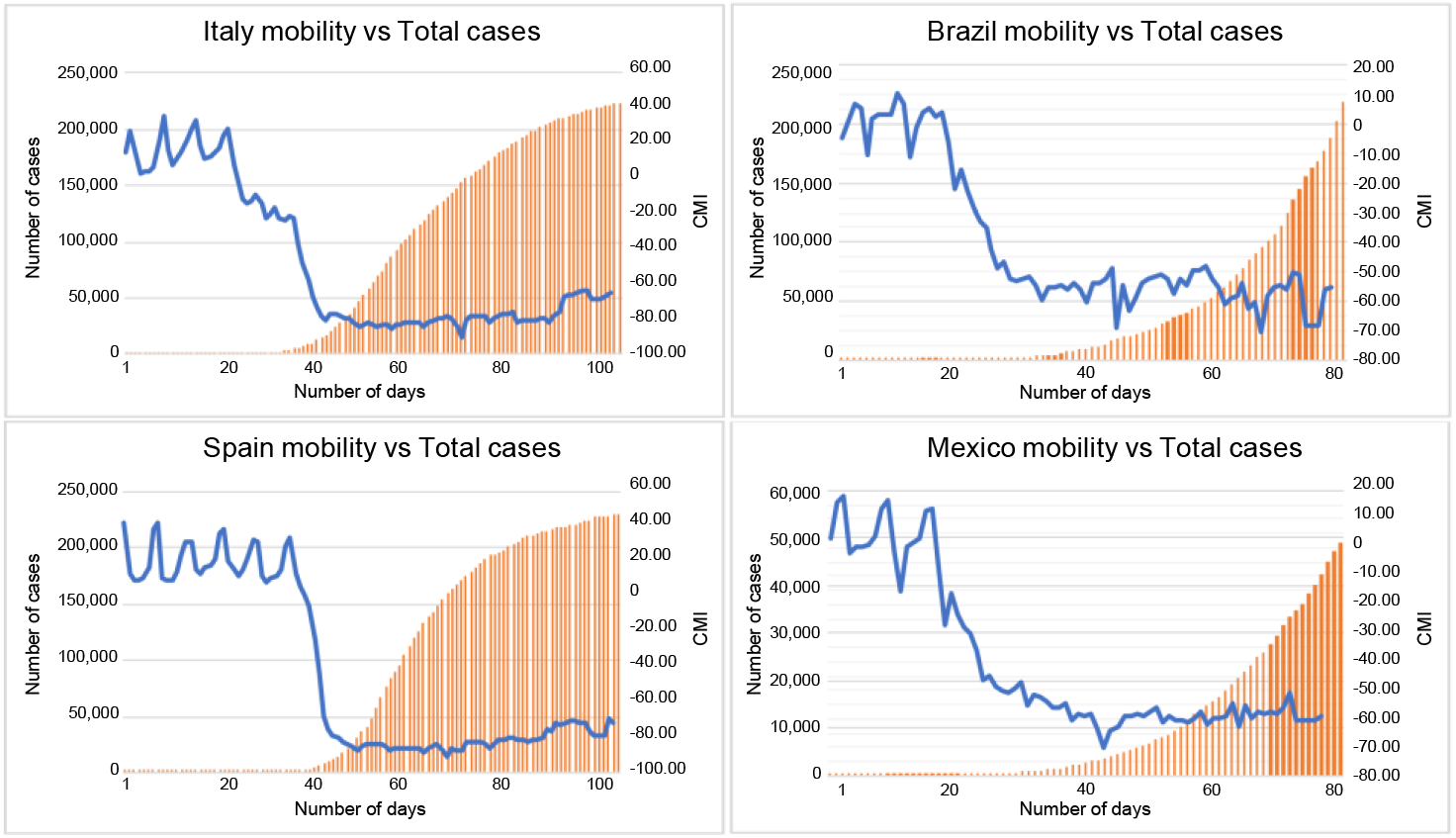
Italy and Spain behavior of number of cases and ***CIM***.

In contrast, countries like Mexico and Brazil, which still show a growing behavior, even both have strong correlations, the difference is in the second 14-day period for number of cases. Mexico had around 500 cases whereas Brazil had almost 2000 cases in the same period of time. It can be expected that mobility reductions in Brazil might be stronger than Mexico ones. Figure 7 also shows the pandemic curves and their ***CIM***.

## 6. Conclusions

In this article it was described the impact on mobility reductions in the public space as a measure to stop the pandemic wave of SARS-COV-2 virus. Also it was discussed mobility reductions as a way to project potential scenarios depending upon a variable called ***CIM*** as a method to monitor cases growth but also to control them. Three scenarios were discussed:

- Light measures, large number of cases, heavy mobility reductions, several impact on economies.
- Moderate measures, manageable number of cases, risk to case growth if not surveillance is applied with potential impact on public health system.
- Strong measures, reduced number of cases, manageable reduction on mobility and constant surveillance of cases to manage mobility recommendations. Public health system is not affected nor local economies.

As discussed, countries like Spain and Italy had to practically stop their economies in order to stop virus spread crashing their public health systems due to a poor surveillance program to monitor virus spread and mobility reductions. 60 days projections for US might anticipate heavy growing by observing their negative correlation and actual number of cases validate the impact on mobility. For Brazil, it can be anticipated that even its strong correlation, initial 30 days case growth will impact future case growth, making mobility measures much stronger.

Four main takeaways are made after all this analysis:

1. The importance of monitor case daily increase for the first 4-6 weeks of the pandemic is crucial to determine reduction in mobility and proper actions.
2. Any 2-digit increase in ***R***_*e*_ will result in heavy case growth if mobility is not reduced as early as possible at levels to at least -20%.
3. ***m***_0_ parameter is determined by two factors: mobility reduction change and phase in the pandemic wave, the earliest, the better. Every 1% reduction in ***m***_0_ is variable depending upon the trend of ***CIM*** and ***R***_*t*_ reductions but is in the range of 0.1 and 0.005 for every 1% of ***CIM***. The earliest identification on case increase will require less mobility reductions.
4. A balance between stronger mobility reduction measures and lighter ones depends on ***R***_*t*_ value and pandemic wave phase (an indirect variable of number of cases as described). ***CIM*** reduction can be in the range of -20% up to -80%.

## Data Availability

All data produced in the present study are available upon reasonable request to the authors

## 7. Author contribution statement

**Morales-Fajardo:** Conceptualization, Methodology, Formal analysis, Investigation, Writing - Original Draft.

**Rodríguez-Arce:** Conceptualization, Methodology, Writing - Original Draft.

## Funding

No external funding.

## Declaration of competing interest

None.

## Notes

### Competing Interest Statement

The authors have declared no competing interest.

### Funding Statement

This study did not receive any funding

